# A Development Framework for Trustworthy Artificial Intelligence in Health with Example Code Pipelines

**DOI:** 10.1101/2024.07.17.24310418

**Authors:** Carlos de-Manuel-Vicente, David Fernández-Narro, Vicent Blanes-Selva, Juan M García-Gómez, Carlos Sáez

## Abstract

Technological trends point to Artificial Intelligence (AI) as a crucial tool in healthcare, but its development must respect human rights and ethical standards to ensure robustness and safety. Despite general good practices are available, health AI developers lack a practical guide to address the construction of trustworthy AI. We introduce a development framework to serve as a reference guideline for the creation of trustworthy AI systems in health. The framework provides an extensible Trustworthy AI matrix that classifies technical methods addressing the EU guideline for Trustworthy AI requirements (privacy and data governance; diversity, non-discrimination and fairness; transparency; and technical robustness and safety) across the different AI lifecycle stages (data preparation; model development, deployment and use, and model management). The matrix is complemented with generic but customizable example code pipelines for the different requirements with state-of-the-art AI techniques using Python. A related checklist is provided to help validate the application of different methods on new problems. The framework is validated using two representative open datasets, and it is provided as Open Source to the scientific and development community. The presented framework provides health AI developers with a theoretical development guideline with practical examples, aiming to ensure the development of robust and safe health AI and Clinical Decision Support Systems. GitHub repository: https://github.com/bdslab-upv/trustworthy-ai

## 1. Introduction

It is widely acknowledged that artificial intelligence (AI) based decision support systems will be of utmost importance to our current and future society, particularly in the health domain. However, the rapid development of this technology has not adequately considered the implications of human rights as set out in the Charter of Fundamental Rights of the European Union (EU Charter) (Charter of Fundamental Rights of the European Union, 2012). The failure to address these declarations could have deleterious consequences for society (Raso et al., 2018), and potentially risk the safety and fundamental rights of millions of patients (Sáez et al., 2024). It results imperative that AI based systems are designed to be trustworthy throughout their entire life cycle.

The European Commission identified three components of Trustworthy AI (TAI): lawful, ethical and robust (European Comission, 2019). Furthermore, the EU outlines four ethical principles to ensure compliance with fundamental rights without hierarchical priority: respect for human autonomy, empowers cognitive, social, and cultural capabilities to maintain self-determination; prevention of harm, safeguards physical and mental integrity, addressing vulnerabilities from power or information asymmetries; fairness, ensures equitable distribution of benefits, prevents bias, discrimination, and requires fair decision-making processes; explainability, promotes transparency in AI processes, varying by application to ensure comprehensibility and accountability for potential errors. The aforementioned ethical principles are addressed through the following TAI requirements, which must be considered throughout the AI system’s life cycle (European Comission, 2019): human agency and oversight, technical robustness and safety, privacy and data governance, transparency, diversity, non-discrimination and fairness, societal and environmental wellbeing, and accountability.

The European Union also exhibits a legal framework constraint as a result of the AI Act (European Parliament, 2024). Nevertheless, the quantitative limits set by the law remain ambiguous. The AI Act is a risk-based proposal. The objective of this initiative is to establish a systematic approach for determining the conformity of high-risk. These steps are based on the requirements proposed in the guidelines. Nevertheless, this regulatory framework does not provide a technical or concrete methodology for achieving trustworthy AI.

Despite the EU’s stipulation that the requirements for trustworthy AI must be met throughout the system’s entire lifecycle, there is currently no consensus on the specific phases that must be addressed. The CRISP-DM framework (Hotz, 2018), used for data mining, includes phases like business understanding, data understanding, data preparation, modeling, evaluation, and deployment. The GenAI lifecycle (Saltz, 2024), for building generative AI applications, defines steps such as problem definition, data investigation, data preparation, development using large language models, evaluation, and deployment. The AWS Well-Architected Framework (Amazon Web Services, 2023), used for developing machine learning projects on AWS, outlines stages including business objective identification, ML problem formulation, data processing, model development, implementation, and monitoring. Lastly, the MLOps approach (Neupane, 2023), aimed at automating and managing the machine learning lifecycle, involves design, model development and operations. Therefore, there is also a need to harmonize and frame well-established AI life-cycle stages with the requirements for trustworthy AI.

Since a number of entities have identified and proposed methods for addressing trustworthy AI (Deloitte, 2022; IBM, 2021) or proposed guidelines for prediction model studies based on artificial intelligence (Collins et al., 2021), despite the significance and relevance of this issue in health, to our knowledge there is currently no available comprehensive methodology classifying open and technical methods to support health AI developers develop and use systems that fully respect all necessary ethical guidelines and requirements.

Consequently, in this work we propose a development framework to serve as a reference guideline for the creation of trustworthy AI systems in health supported with example code pipelines comprising technical methods that may be used to fulfil the trustworthiness requirements and achieve a TAI in the health sector. The framework is validated using two open healthcare datasets, and all codes and guidelines are published as Open Source.

## 2. Materials and methods

### 2.1. Workflow

*Figure 1* describes the workflow of this work, whose aim is to provide a clear and systematic framework to develop a reliable and generalizable AI solution. As such, the methodology starts by establishing the requirements for TAI to be addressed, alongside the harmonization of AI lifecycle stages. Next, for each TAI requirement state of the art technical methods are classified and organized throughout the AI lifecycle stages, which are then compiled into specific code notebooks. The components for each requirement and AI stage are sorted in a trustworthy AI matrix, which helps relate and navigate through the proposed concepts. Next, a related checklist of accomplishments is proposed while the notebooks are validated with two open datasets.

**Figure 1.**
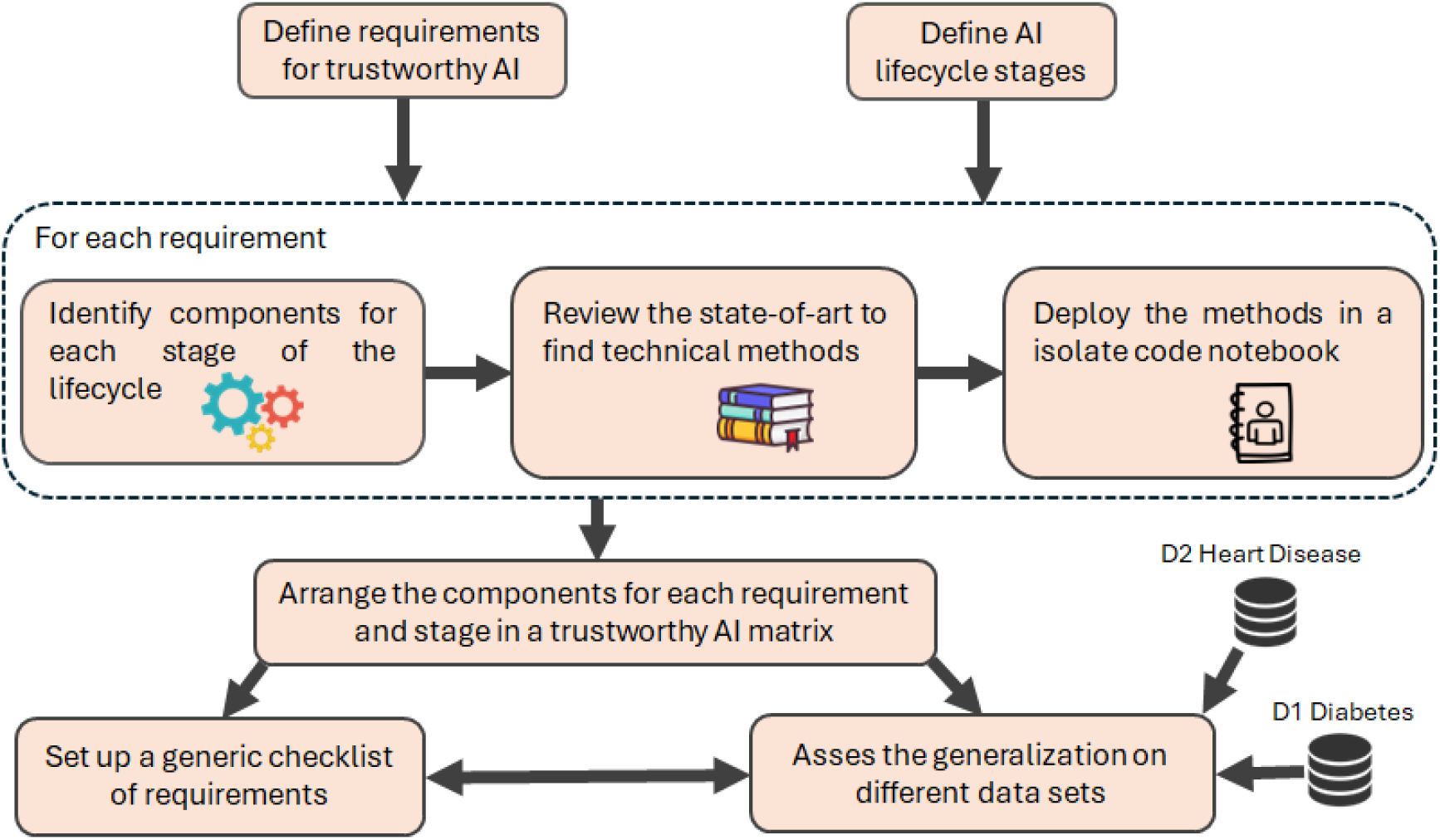
Flowchart of the work methodology.

**Figure 2.**
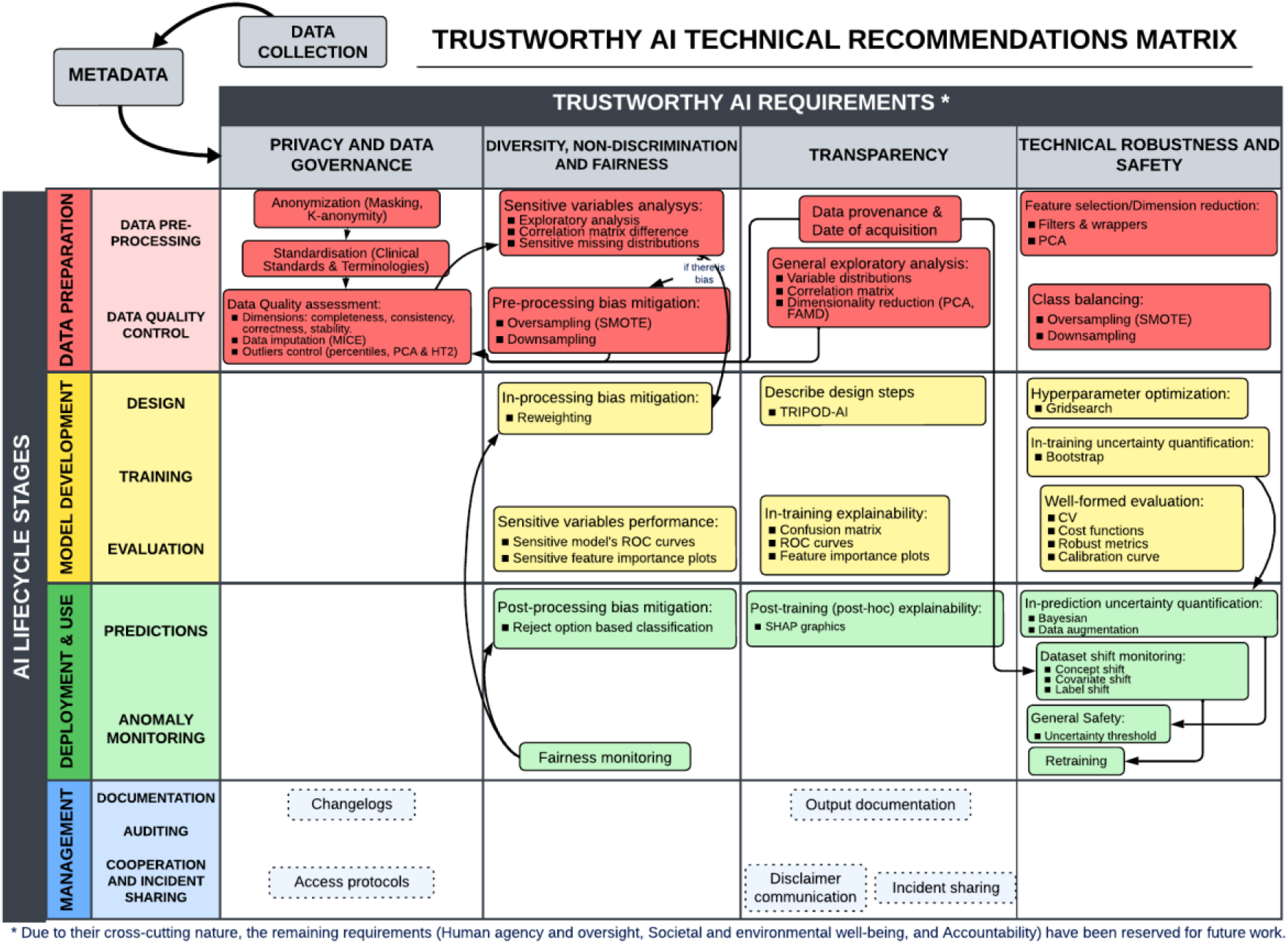
Requirements matrix for trustworthy AI (columns) by lifecycle stage (rows). The boxes represent the components necessary to fulfil each requirement. Technical methods are indicated by items within the boxes. Discontinued boxes indicate that the components are not strictly related to provided technical methods. Arrows indicate potential dependencies or workflows between the components.

Within AI and specifically machine learning, the predictive branch includes classification problems with categorical or discrete outputs (e.g., readmission or no readmission) and regression problems with numerical or continuous outputs (e.g., number of months of patient survival). Currently, the technical methods proposed in study focus on classification problems, although regression problems could be addressed in future work by extending the current guidelines.

### 2.2. Tools

The study integrates Python notebooks in Jupyter Notebook (versions 3.12.1 and 7.0.7) within Visual Studio Code (VS Code), effectively combining code with text to demonstrate the development of reliable AI models. Python’s versatility and robustness, alongside its rich library support for data analysis and machine learning, made it a leading choice in this work.

The preference for notebooks over purely code-based files underscores their pivotal role in providing extensive, clear tutorials with executable AI development processes. This structured approach, akin to literate programming (Knuth, 1984), aids comprehension and guides users through workflows, preventing confusion that might arise from less clear, context-lacking code files. Furthermore, the study employs machine learning algorithms such as Random Forest and Naive Bayes from the scikit-learn Python library. Random Forest constructs multiple decision trees and aggregates their predictions, offering robustness against overfitting and suitability for complex datasets with diverse features and classes (Shanthakumari et al., 2022). Naive Bayes, based on Bayesian theorem, assumes feature independence (Sureskumar, 2017) and is implemented here using Python’s ‘GaussianNB’ and ‘CategoricalNB’ models.

### 2.3. Datasets

The proposed framework is primarily validated using the “Diabetes 130-US Hospitals for Years 1999-2008” dataset (John Clore, 2014). It is constructed using data from 101 768 encounters of diabetic patients with a classification problem. The goal is to predict whether there will be a readmission within <30 days, >30 days, or no readmission after discharge. It includes numerical and categorical features from different nature, such as demographic or diagnosis.

In order to verify the generalization of the AI model’s performance across different healthcare conditions, the framework is also validated using the “Heart Disease Set”(Ali et al., 2021; Alizadehsani et al., 2019). This dataset is renowned for its application as a benchmark in health AI and dates back to 1988, combining data from Cleveland, Hungary, Switzerland, and Long Beach. It consists of 11 features and 1190 encounters, with the target label indicating the presence of heart disease (1 for presence, 0 for absence). The dataset includes 5 numerical and 6 categorical variables, which notably have fewer categories compared to those in the Diabetes Set, making it a suitable complement to validate the proposed AI model’s reliability across diverse medical datasets.

## 3. Results

Next, we describe the results including the proposed Trustworthy AI matrix, the validation on the two datasets, and the proposed checklist of recommendations. In the spirit of open science, all the following results, as well as the source code, are publicly available on the GitHub repository: https://github.com/bdslab-upv/trustworthy-ai.

### 3.1. Trustworthy AI matrix

The study introduces a matrix for trustworthy AI to guide developers in constructing reliable AI models by correlating lifecycle requirements, aligned with the European Commission’s TAI guidelines (European Comission, 2019). After reviewing methodologies, we propose lifecycle stages as follows: Data Preparation involves data pre-processing for cleaning, transformation, feature selection, and quality control. Model Development includes designing, training, and validating models, selecting types and parameters while adhering to ethical and technical robustness principles. Deployment and Use assesses real-world model implementation, monitors ongoing performance, and adjusts as needed. Management involves auditing results, documenting model details, and conducting regular audits for operational integrity and stakeholder collaboration.

Preceding the matrix requirements, there is a Metadata step for dataset conditioning and introducing additional information transversal to the principles. Though the EU dictates no hierarchy (European Comission, 2019), we suggest column-based resolution: beginning with Data Privacy and Governance to ensure appropriate material handling; Diversity, non-Discrimination and Fairness to prevent or quantify biases during training; Transparency for comprehensive data understanding and supplementary tool development; and lastly, Robustness and Security for model performance and uncertainty management assurance. Addressing the transversality of the remaining requisites, integrating them within established categories ensures a comprehensive approach to AI system development aligned with ethical and regulatory frameworks.

The remaining three, Human Agency and Supervision, Social and Environmental Well-being, and Accountability, for their indirect technical resolution and close transversality regarding the former, are proposed to be included in future work. Nevertheless, although the scope of these requirements is not defined, they are partially addressed in other components. Human Agency and Supervision could align within Diversity, Non-Discrimination, and Justice, due to their fundamental rights implications. Accountability relates to Transparency’s explainability and auditability in Robustness and Security, warning of significant performance declines prompting retraining.

A further significant outcome is the creation of pipelines as a generic implementation tool. In accordance with the methodologies proposed in the TAI Matrix, a comprehensive notebook has been constructed, incorporating code and detailed explanatory text for each specified requirement. To facilitate comprehension of the study, the following section presents an overview of some methodologies employed in each notebook, along with a discussion of their assessment on diverse health data sets. Emphasizing the use of external functions or pipelines to streamline code complexity, the framework includes a standalone implementation of the Naive Bayes mixed model and a ‘handleData’ class designed to handle data transformations, such as encoding or feature grouping.

#### 3.1.1. Data Collection and Metadata

As an antecedent step to the TAI matrix, data collection encompasses all code for properly obtaining data in an analysable format, such as in a tabular format. The transition from this stage to the guide necessitates an intermediate phase for the preparation of required metadata. The objective of this phase is to specifically formatting the data for the developed pipeline, including additional information that is crucial to meet various requirements.

Once metadata is compiled and adapted to the dataset, it is exported to a JSON file for easy access across requirements, facilitating the development of trustworthy AI, starting with Data Privacy and Governance.

#### 3.1.2. Privacy and Data Governance

The Privacy and Data Governance section is concerned with the assurance of data quality, integrity, and compliance with data privacy regulations through the application of anonymization techniques. These include k-anonymity (Murthy et al., 2019; Olatunji et al., 2022), which serves to prevent the identification of individuals, as well as standardization (e.g., OMOP-CDM for healthcare (OHDSI, 2024)), and data quality control (e.g., completeness, consistency, correction) (Sáez et al., 2012, 2024). The completeness and correction dimensions are addressed technically. Outliers are treated with in two ways: univariate outliers are treated with using the 95th percentile, while multivariate outliers are treated with using principal component analysis (PCA) with Hotelling’s T-squared test (HT2) (Taskesen, 2023), with a significance level of α=5%. Furthermore, all forms of missing data (MCAR, MAR, MNAR) are addressed using MICE imputation methods (Stavseth et al., 2019), specifically linear regression and KNN imputers. The management of data post-implementation would include the maintenance of change logs, access protocols, and the implementation of rigorous data handling protocols to restrict access to authorized personnel only.

#### 3.1.3. Diversity, non-discrimination and fairness

This section emphasizes inclusivity and the protection of fundamental rights throughout the AI lifecycle. In Data Preparation, there’s a focus on conducting sensitive subgroup analyses to meticulously scrutinize data for any disparities linked to sensitive variables. A comparison of different methods for bias mitigation is offered. Oversampling techniques such as Synthetic Minority Oversampling Technique (SMOTE) (Krasanakis et al., 2018) are employed to mitigate biases pre-training. This involves modifying the previous data distribution in order to achieve a more balanced dataset. Moving to Model Development, reweighting, which assigns weights to categories based on their prevalence in the dataset, is implemented to counteract biases during training. Additionally, performance evaluations across sensitive subgroups help identify and rectify any disparities that may affect model fairness.

During Implementation and Use, efforts continue with post-processing techniques such as Reject Option Based Classification (Kamiran et al., 2018), designed to handle uncertain classifications conservatively, particularly in cases where disadvantaged groups are involved. The section emphasizes using color palettes like “viridis” or “colorblind” configurations (Michael Waskom, 2012) to improve accessibility for individuals with color vision deficiencies, aligning with efforts to promote equity and justice in AI applications.

#### 3.1.4. Transparency

This requirement aims to achieve a thorough understanding of the data, model, and predictions through exploration, explanation, and documentation processes. Initial data preparation involves extracting lineage information from metadata to contextualize data appropriately. General exploratory analysis aims to uncover correlations between variables and the predicted class, utilizing techniques such as correlation matrices for numerical data and bar charts for categorical data to assess class proportions per category. During the development of the model, it is of the utmost importance to document the rationale behind the selection of the model, the adoption of the parameters, and the training and validation methods. This can be achieved by utilizing frameworks such as TRIPOD (Collins et al., 2021). Furthermore, explainability graphics, such as confusion matrices and ROC curves, aid in the understanding of the model decisions. Once the training process has been completed, methods such as SHAP (SHapley Additive exPlanations)(Albahri et al., 2023; Molnar, 2021) are employed for model interpretation, which highlights the feature contributions to the predictions. Management includes the documentation of predictive model outcomes and the provision of a disclaimer to acknowledge the characteristics and limitations of the model, with the objective of facilitating incident reporting in order to address unforeseen risks to end-users or stakeholders.

#### 3.1.5. Technical Robustness and Safety

Finally, technical robustness and security aim to prevent or minimize unintended harm by using comprehensive methods to improve model performance and accurately assess its behaviour throughout its lifecycle. In the context of data preparation, it is of paramount importance to implement critical steps such as feature selection or dimensionality reduction in order to mitigate the so-called “curse of dimensionality” (Karanam, 2021), which could affect model robustness. Furthermore, it is of paramount importance to address class imbalances through the use of techniques such as SMOTE (Elreedy & Atiya, 2019). In the context of model development, the focus is on hyperparameter optimization through techniques such as grid search, which ensures the optimal selection of parameters for model performance. In addition, the optimal probability threshold for the model must be identified in consideration of the trade-off between sensitivity and specificity for each context.

The quantification of uncertainty during training is achieved through the utilization of confidence intervals, while robust evaluation methods such as cross-validation are employed to ensure the reliability and accuracy of model predictions. In the implementation phase, strategies for uncertainty quantification involve the utilization of confidence intervals derived from the training process (Bayesian methods) and data augmentation methods (Abdar et al., 2021). These methods compare the consistency of multiple methods applied to the same sample, as well as a single method applied to multiple times perturbed data.

Furthermore, the monitoring of dataset shifts once a model is put into production enables the evaluation of potential data variability over time or across different contexts (Sáez et al., 2012). This enables the adaptation of models through retraining strategies, thereby maintaining accuracy and security in the face of changing distributions. These measures collectively aim to ensure the model’s stability and reliability in real-world applications, thereby minimizing the risk of inaccurate predictions and ensuring robust performance across diverse scenarios.

### 3.2. Check-list of recommendations

In addition to the previously established Trustworthy AI Matrix, a generic checklist has been developed to facilitate the implementation of trustworthy AI in accordance with EU requirements. This checklist consolidates all aspects necessary to achieve the aforementioned goal. *Table 1* presents a comprehensive checklist that encompasses all addressed requirements, along with the supporting technical components and methods. It is designed to be expandable, allowing for the incorporation of new technical methods to address unaddressed requirements or enhance existing ones, such as preventive attack methods.

**Table 1.**
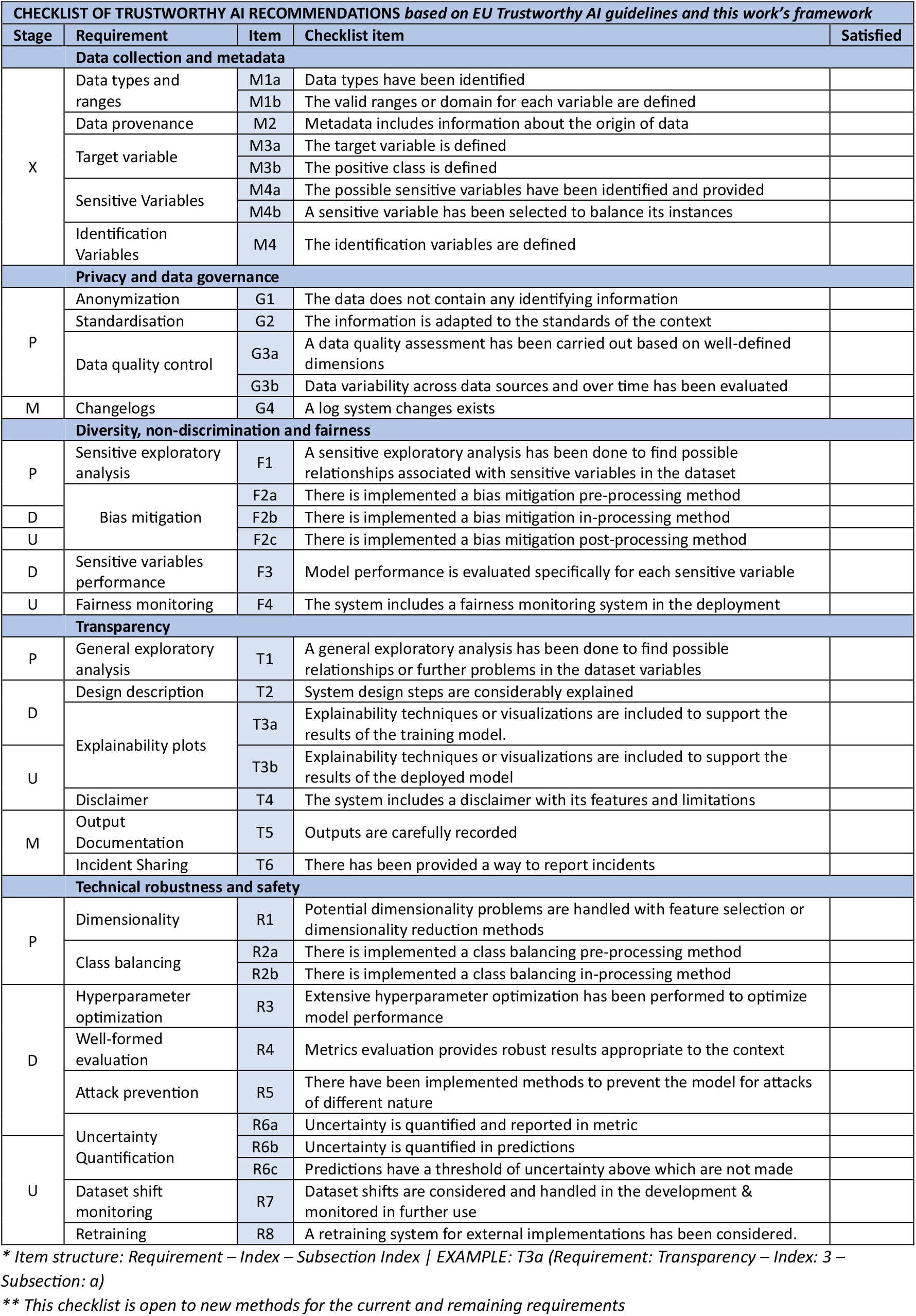
Checklist of recommendations for trustworthy AI according to requirement and lifecycle (P: Data preparation, D: Model development, U: Deployment & Use, M: Management, X: Not part of the life cycle)

### 3.3. Evaluation of datasets

In this section we describe some of the results of validating the TAI framework with the two open datasets. Full results are described in the project GitHub repository.

The metadata parameters for each dataset, as a stage previous to entering the TAI matrix, are described in *Table 2*.

**Table 2.** Initial metadata for diabetes and heart disease datasets. Names of variables are single quoted. Descriptions for the datasets are provided in section 2.3.

|  | DIABETES DATASET | HEART DISEASE DATASET |
| --- | --- | --- |
| **Dataset** | 'dataset_diabetes_simplified.csv' | 'dataset_heart_disease_full.xlsx' |
| **Output** | 'readmitted' | 'target' |
| **Positive class** | '<30' | '1' |
| **Identifying features** | 'encounter_id' y 'patient_nbr' | ' ' |
| **Sensitive features** | 'race' y 'gender' | 'sex' |
| **Balancing feature** | 'race' | 'sex' |
| **Data provenance** | ["A Health Facts database that represents 10 years (1999-2008) of clinical care at 130 hospitals in United States.", 'admission_type'] | ["The dataset consists of 1190 records of patients from US and Hungary", ' '] |
| **Acquisition date** | ["Empty", ' '] | ["Empty", ' '] |
| **Variable types** | ▪ race: categorical<br>▪ gender: categorical<br>▪ age: categorical<br>▪ weight: categorical<br>▪ admission_type: categorical<br>▪ discharge_disposition: categorical<br>▪ admission_source: categorical<br>▪ time_in_hospital: numerical<br>▪ payer_code: categorical<br>▪ medical_specialty: categorical<br>▪ num_lab_procedures: numerical<br>▪ num_procedures: numerical<br>▪ number_outpatient: numerical<br>▪ number_emergency: numerical<br>▪ number_inpatient: numerical<br>▪ diag_1: categorical<br>▪ diag_2: categorical<br>▪ diag_3: categorical<br>▪ number_diagnoses: numerical<br>▪ change: categorical<br>▪ diabetesMed: categorical | ▪ age: numerical<br>▪ sex: categorical<br>▪ chest_pain_type: categorical<br>▪ resting_bp_s: numerical<br>▪ cholesterol: numerical<br>▪ fasting_blood_sugar: categorical<br>▪ resting_ecg: categorical<br>▪ max_heart_rate: numerical<br>▪ exercise_angina: categorical<br>▪ oldpeak: numerical<br>▪ ST slope: categorical |

Regarding the Diversity, non-discrimination and fairness requirement, *Figure 3* illustrates the presence of imbalanced sensitive features in both datasets, along with the implementation of various solutions. With respect to the Diabetes dataset, it is feasible to address the issue of racial imbalance by examining the relative importance of each variable in models trained with different racial groups. In contrast, the Heart Disease dataset has been subjected to a comparison of three techniques to address the issue of sex imbalance. The first is to take no action, which may be considered a simple approach. The second is to mitigate bias pre-training (oversampling), and the third is to address the issue during training (reweighting).

**Figure 3.**
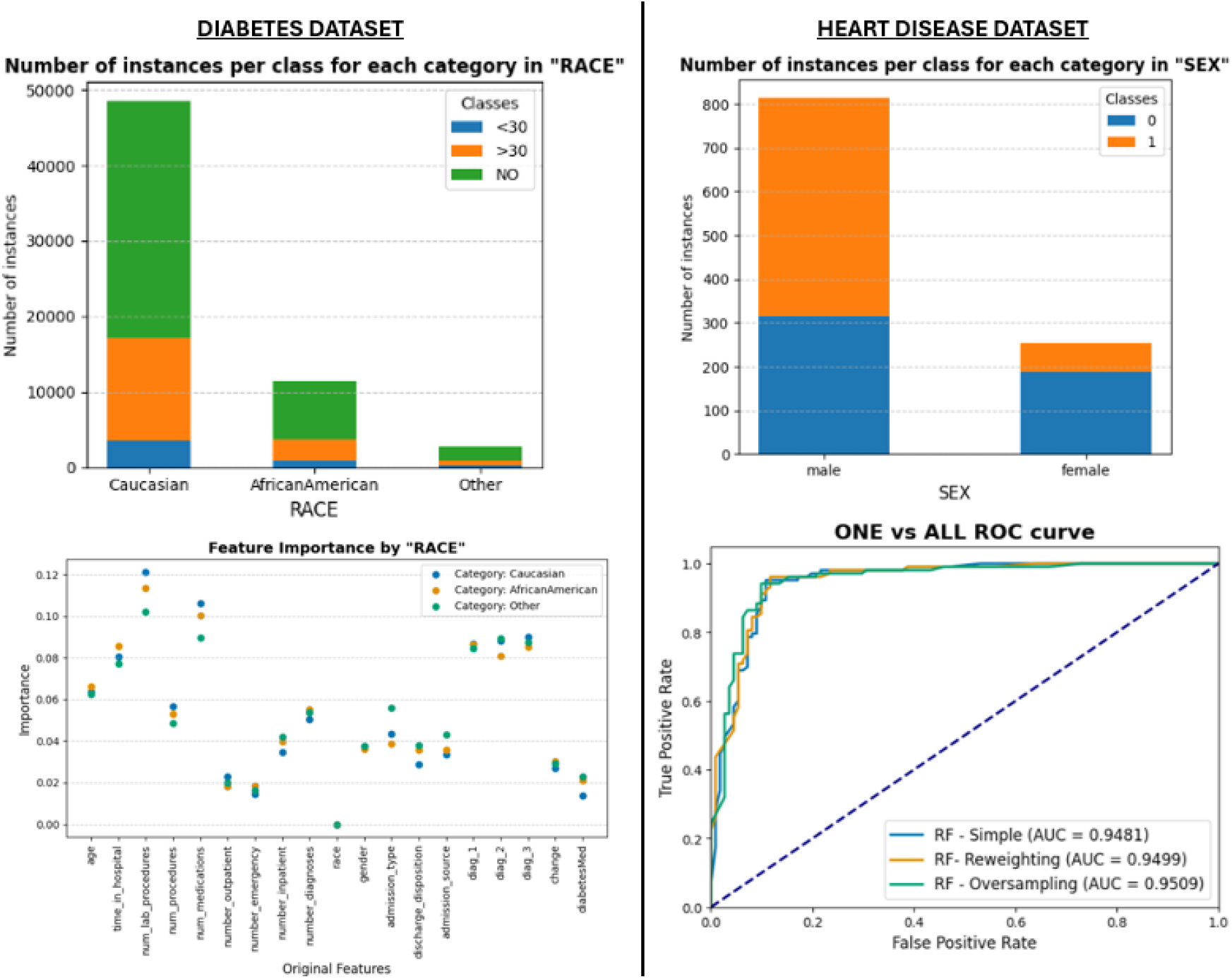
Selection of the Diversity, non-discrimination and fairness notebook results for both datasets.

Regarding the Transparency requirement, for the Diabetes dataset, *Figure 4* illustrates the in-training feature importance, which could enhance explainability and could be employed in feature selection by applying importance thresholds. For the Heart Disease dataset, a bivariate plot is presented for general exploratory analysis.

**Figure 4.**
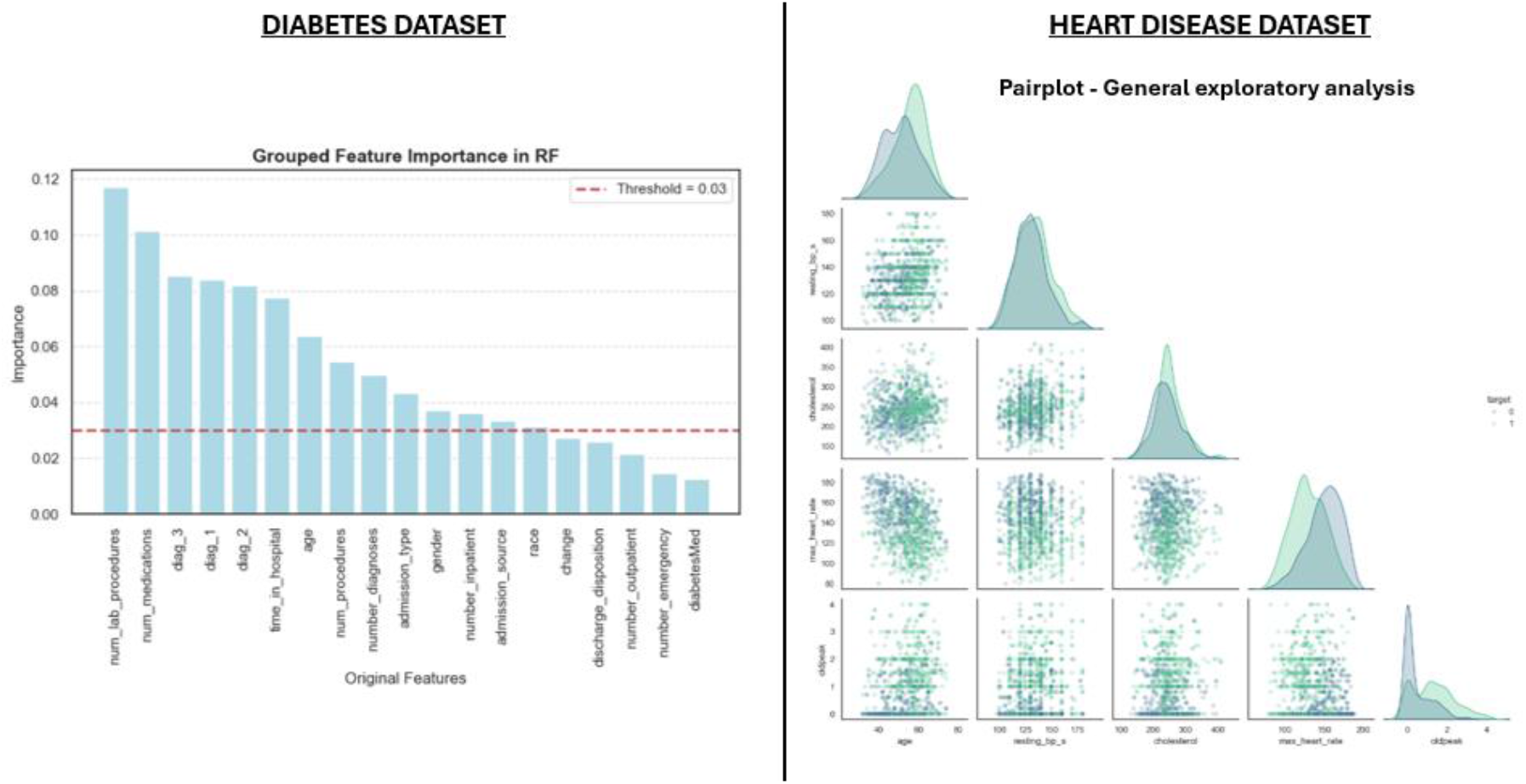
Selection of the Transparency notebook results for both datasets.

Regarding the Technical robustness and safety requirement, *Figure 5* illustrates a selection of the results for both datasets. With regard to robustness, the figure depicts the ROC curve resulting from the optimization of the training process in Diabetes dataset, whereas the plot for Heart Disease shows its calibration curve. In addition, the uncertainty in predictions is quantified through the use of ten models for the prediction of an instance in Diabetes dataset and one model for the prediction of one hundred perturbed instances in Heart Disease.

**Figure 5.**
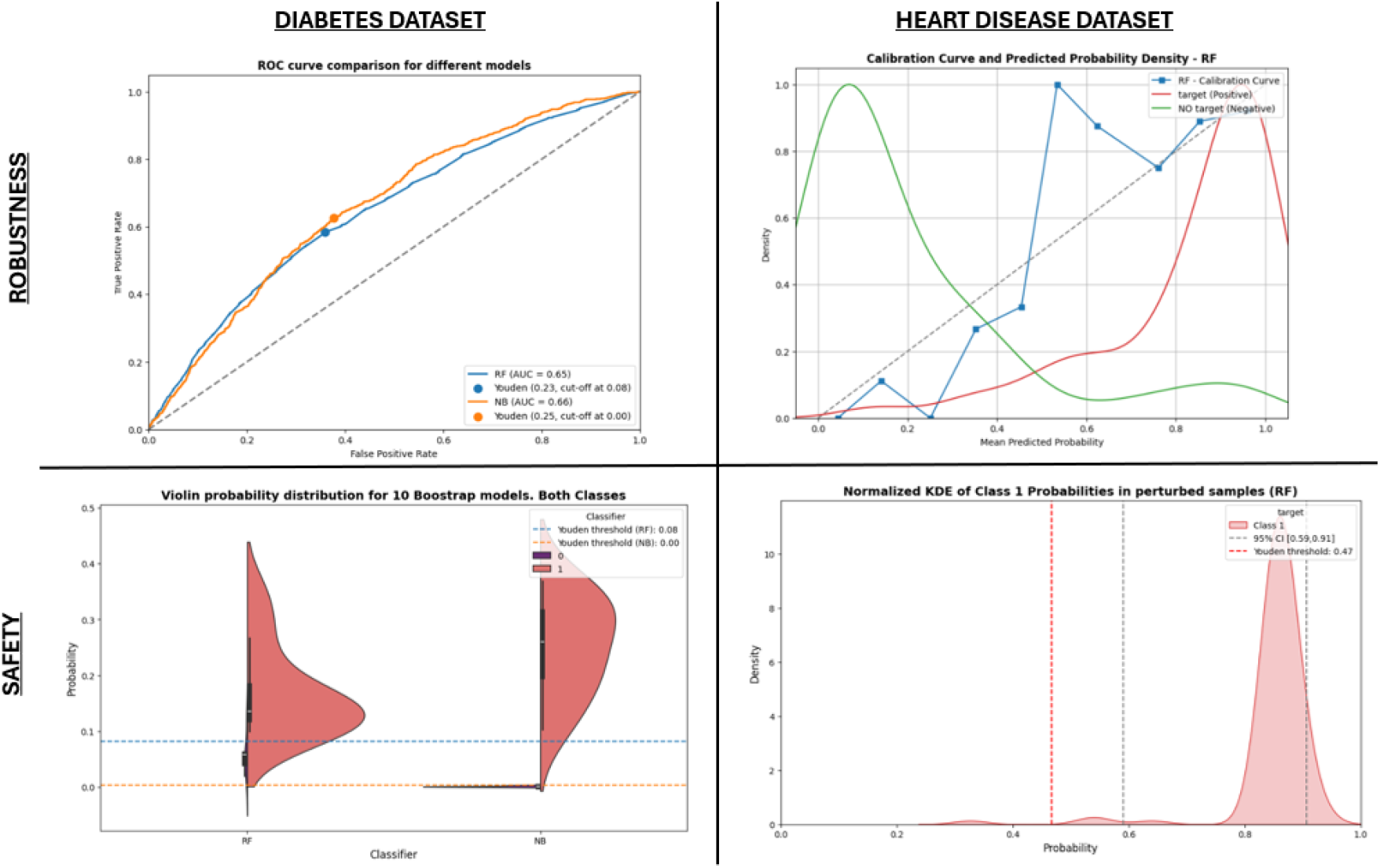
Selection of the Technical Robustness and Safety notebook results for both datasets.

## 4. Discussion

This work presents the development of a trustworthy AI framework tailored for healthcare environments, in alignment with European guidelines such as the European Commission’s guidelines on trustworthy AI (European Comission, 2019) and the AI Act (European Parliament, 2024), and aims to provide a step further towards achieving resilient AI in health (Sáez et al., 2024). The methodology provides a robust framework for data engineers and other specialists to construct trustworthy AI systems, demonstrating high adaptability to various datasets. The practical value of this approach lies in its provision of executable code and hands-on implementation strategies. While the system performs optimally with ideal datasets such as the “Heart Disease” dataset and adapts to real-world complexities as seen with the “Diabetes” dataset, it remains a semi-automated pipeline that can be fine-tuned for specific implementation contexts to ensure maximum efficiency and reliability.

This work is situated at the vanguard of the most advanced techniques for developing trustworthy AI, integrating methodologies that address all EU-established requirements for health AI. Regarding technical methods, this approach differs from others, such as those of Deloitte (Deloitte, 2022) and IBM (IBM, 2021), which adopt a segmented methodology addressing trustworthy AI components individually. Other AI guidelines, such as TRIPOD-AI (Collins et al., 2021), not only focus uniquely on specific stages, such as model development or validation, but also do not address trustworthiness. In contrast, the current methodology encompasses the entire trustworthy AI lifecycle, from data collection to implementation and management. This addresses a significant gap in the field.

Future work will concentrate on increasing the full automation of the pipeline, enhancing its capabilities with deep learning techniques for broader data generalization, such as using imaging or open text data, and addressing additional EU requirements such as Human Agency and Oversight, Social and Environmental Well-being, and Accountability. The expansion of metadata in accordance with the European Health Data Space (EHDS) (European Commission, 2022; European Union Agency for Fundamental Rights, 2019) will facilitate the standardization and interoperability of data for health AI, thereby enhancing transparency and reproducibility. Further validation with a larger user base will enable the methodology to be refined based on practical feedback.

## 5. Conclusions

The proposed framework establishes a pipeline for the creation of trustworthy AI in healthcare, in alignment with EU principles and requirements pertaining to privacy and data governance, diversity, non-discrimination and fairness, transparency, and technical robustness and safety. We provide a comprehensive methodological guideline that details the necessary actions throughout the AI lifecycle. Additionally, a matrix was developed as a conceptual map that linked methods with lifecycle stages and trustworthy requirements. The creation of dynamic notebooks for each reliability principle, which are available as open-source code, offers practical tools for the development of trustworthy AI systems. The pipeline demonstrated generalisability across various healthcare datasets, although some human supervision is still required, and three additional EU requirements can be further formalized in the matrix. In conclusion, this work provides a robust foundation for the development of trustworthy AI in healthcare. It offers a practical and adaptable framework that can be expanded and refined to provide optimal solutions in specific contexts aiming to ensure the development of robust and safe health AI and Clinical Decision Support Systems.

## Data Availability

All data used are openly available at the UCI Machine Learning repository.

https://github.com/bdslab-upv/trustworthy-ai/

## Acknowledgements

This work has been funded by the Spanish ‘Ministerio de Educación, Beca de colaboración (2023/12/00002), Inteligencia artificial confiable en biomedicina’, and ‘Agencia Estatal de Investigación, Proyectos de Generación de Conocimiento (PID2022-138636OA-I00), KINEMAI’.

